# The Psychedelic Media Exposure Questionnaire: The development and validation of a new scale assessing psychedelic-related media exposure

**DOI:** 10.1101/2025.06.30.25330536

**Authors:** Audrey Evers, Chris Kelly, Tehseen Noorani, Shayla Love, Boris D. Heifets

## Abstract

**Rationale:** A reliable and valid instrument is needed to measure exposure to psychedelic-related media to examine the impact of media and expectations on outcomes in psychedelic clinical trials.

**Objectives:** Our aim was to explore the psychometric properties of the Psychedelic Media Exposure Questionnaire (PMEQ) and to validate the scale.

**Methods:** First, items were developed by a team of experts, reviewed by external expert collaborators, and revised. Psychometric validation of the PMEQ was carried out using data from two participant samples. The measure was administered to a registry of patients who had agreed to be contacted for research opportunities at Stanford Health Care, of whom 472 completed the measure (Study 1). Exploratory factor analysis was performed, and internal consistency was tested. The measure was then administered to a sample of social media users via various online platforms to further validate the scale, of whom 159 completed the measure (Study 2). Confirmatory factor analysis was performed, and internal consistency was tested.

**Results:** Factor analysis of the PMEQ revealed four main factors with good internal consistency in Study 1 (*ω* =.83 and *α* =.84) and Study 2 (*ω* =.86 and *α* =.85). The four underlying factors are positive attitude toward psychedelics, assessment of the emblematic media piece, engagement with media, and negative attitude toward psychedelics.

**Conclusions:** The PMEQ is a new measure of psychedelic-related media exposure with acceptable psychometric properties. This scale will contribute to our understanding of media bias and patient expectations in both clinical research and real-world settings where psychedelics treatments are used.

## Introduction

*“Psychedelic drugs…are heavily influenced by context, such as a person’s mindset, environment, and beliefs. In other words, people’s expectations about what happens on psychedelic drugs might play a role in what they experience. These problems have haunted the study of psychedelics since the first wave of research in the 1950s…” - (Love, 2023)*

The “Pollan Effect” (Noorani, 2020) and the “Michael Pollan Effect” (Carpenter, 2020) were coined the same year in reference to Michael Pollan’s popular book “How to Change Your Mind” and have been used to highlight the potential impact of media hype on patient outcomes, clinical trial enrollment, and public attention, following Pollan’s landmark publication that popularized the resurgence of psychedelic research (Aday et al., 2022; Pollan, 2018). This “psychedelic renaissance” followed an abrupt halt in the nascent field of psychedelic medicine in the 1970s; over the past decade, a more favorable regulatory environment has allowed research to resume for psychedelic treatment and psychedelic-assisted psychotherapy in a broad range of psychiatric indications, including posttraumatic stress disorder (PTSD), major depressive disorder (MDD), and substance use disorders (Luoma et al., 2020; Mitchell & Anderson, 2024; Reiff et al., 2021). This surge in research has generated an abundance of largely positive media coverage from newspapers, magazines, podcasts, and more. Critical voices in academia (Johnson, 2021) and popular media (Love, 2022) have sounded alarms about over-exuberant expectations, echoing “hype bubbles” about previous treatments such as stem cell interventions, marijuana for anxiety or Parkinson’s disease, and melatonin for sleep, which have left clinicians concerned about misleading medical information in the media (Kamenova & Caulfield, 2015; Robledo & Jankovic, 2017).

In a variety of clinical trials, patients’ expectations have been shown to contribute to treatment efficacy (e.g., deep brain stimulation for Parkinson’s disease; Mameli et al., 2023). Uncontrolled expectations about treatment outcomes may be particularly confounding in the setting of psychedelic clinical trials, where the challenge of adequately masking study group assignments can create unbalanced expectations between treatment groups (Aday et al., 2022; Hall & Humphreys, 2022; Reiff et al., 2021; Schatzberg, 2020; Szigeti et al., 2024; Szigeti & Heifets, 2024; van Elk & Fried, 2023). Additionally, as seen in other hype bubbles, inflated expectations may undermine patient trust and satisfaction when treatment outcomes fail to align with media portrayals (Wilkinson & Sanacora, 2017; Yaden & Griffiths, 2022). Media hype likely contributes to treatment expectations (Aday et al., 2022; Hall & Humphreys, 2022; Noorani et al., 2023; Yaden et al., 2022), but this generally held suspicion about the contribution of media (i.e., newspapers, web-based articles, podcasts) to treatment expectations has not been formally evaluated with respect to the extent or quality of those media-informed beliefs about psychedelics. Treatment decision-making by clinicians and patients is influenced by numerous factors such as education, experience, comfort, and sociodemographic positioning, and amongst these, we propose a deeper consideration of media influences. Through access to media and academic sources, patients have exposure to a variety of content that contribute to their treatment attitude and compliance, which may impact preference for clinical trial enrollment and unaccounted for variance in research samples.

Further, media representations change and develop over time. While it appears that media attention regarding psychedelics has been largely positive in recent years, there may be a shift in media sentiment as the “hype bubble” dies down or even bursts, as suggested by Yaden and colleagues (2022). As discussed by Noorani and colleagues (2023), the changing social zeitgeist may create an unusual situation wherein the efficacy of an intervention becomes unstable. Openly documenting the social and cultural milieu in which clinical trial results exist becomes imperative in this dynamic climate (Noorani et al., 2023).

We sought to develop and investigate the factor structure, reliability, and validity of a new questionnaire, the Psychedelic Media Exposure Questionnaire (PMEQ), in two samples: (1) a Stanford Health Care database of patients willing to be contacted for research and (2) individuals on various online platforms (i.e., Reddit, X, Facebook).

### General Methods

All procedures were reviewed and approved by the Stanford University Institutional Review Board (IRB #72592). Data collection occurred from December 2023 - April 2024. All participants were provided with electronic informed consent, which presented them with the study’s purpose, procedures, rights, and any risks or benefits. Consent was obtained by participants acknowledging their understanding and agreement before proceeding with the survey.

This study was carried out in two phases: an item generation and scale development process (Phase 1), followed by two psychometric validation studies (Phase 2).

### Participants (Phase 2)

Psychometric validation was completed in two independent participant samples (Study 1 and Study 2). Participants in both studies were required to meet the inclusion criteria of being over 18 years old and having a proficient understanding of English. Study 1 participants were patients who had given prior consent for research contact, maintained in a database by the Stanford Pain Clinic Collaborative Health Outcomes Information Registry (CHOIR; Callahan et al., 2023). Following completion of Study 1, we sought to recruit a more diverse sample to assess the generalizability of the scale. Study 2 consisted of non-clinical volunteers gathered via online platforms (i.e., Reddit, X, Facebook). Recruitment was carried out via email for Study 1 and through online methods for Study 2. All data was collected online via Research Electronic Data Capture (REDCap). In the section below, we provide details relevant to all survey development and validation phases.

## Data analysis

Descriptive statistics for both studies are presented as frequencies (and %). Statistical analyses were conducted using the IBM Statistical Package for Social Sciences (SPSS), version 29.0.1.0. and JASP version 0.18.3.

### Phase 1: Item generation and scale development

Five professionals with expertise in psychedelics from clinical psychiatry, philosophy, clinical psychology, psychedelic research, and journalism backgrounds collaborated to identify specific constructs and develop an initial item pool for the scale. Four initial consultation meetings included development, feedback, and expansion of the scale. The group aimed to measure the following constructs: quantity of media exposure, positive or negative attitude of the media, and engagement with the media.

The first section of the measure (**Fig. 1**; Questions 1-5) assesses exposure to psychedelic-related media and engagement with psychedelic-related media in the previous six-month period. These questions are time-specific to improve memory recall and better assess the target construct (Walentynowicz et al., 2018). In the following section (Question 6), participants are asked, in an open-ended format, to identify a singular piece of psychedelic-related media that has shaped their perspective most. Then, participants are asked (Questions 6a-6h) questions related to their selected piece of media that assessed the attitude of, authority of, and engagement with that piece. This structure replicated techniques to improve assessment quality (Kashdan et al., 2020) used for other psychological measures. Optional open-ended questions were included: “In what ways has the piece of media you selected shaped your thinking?” and “What important media narratives are missing from the piece of media you selected above?” These qualitative data techniques were included to provide the opportunity for alternative data analytic procedures.

**Figure 1.**
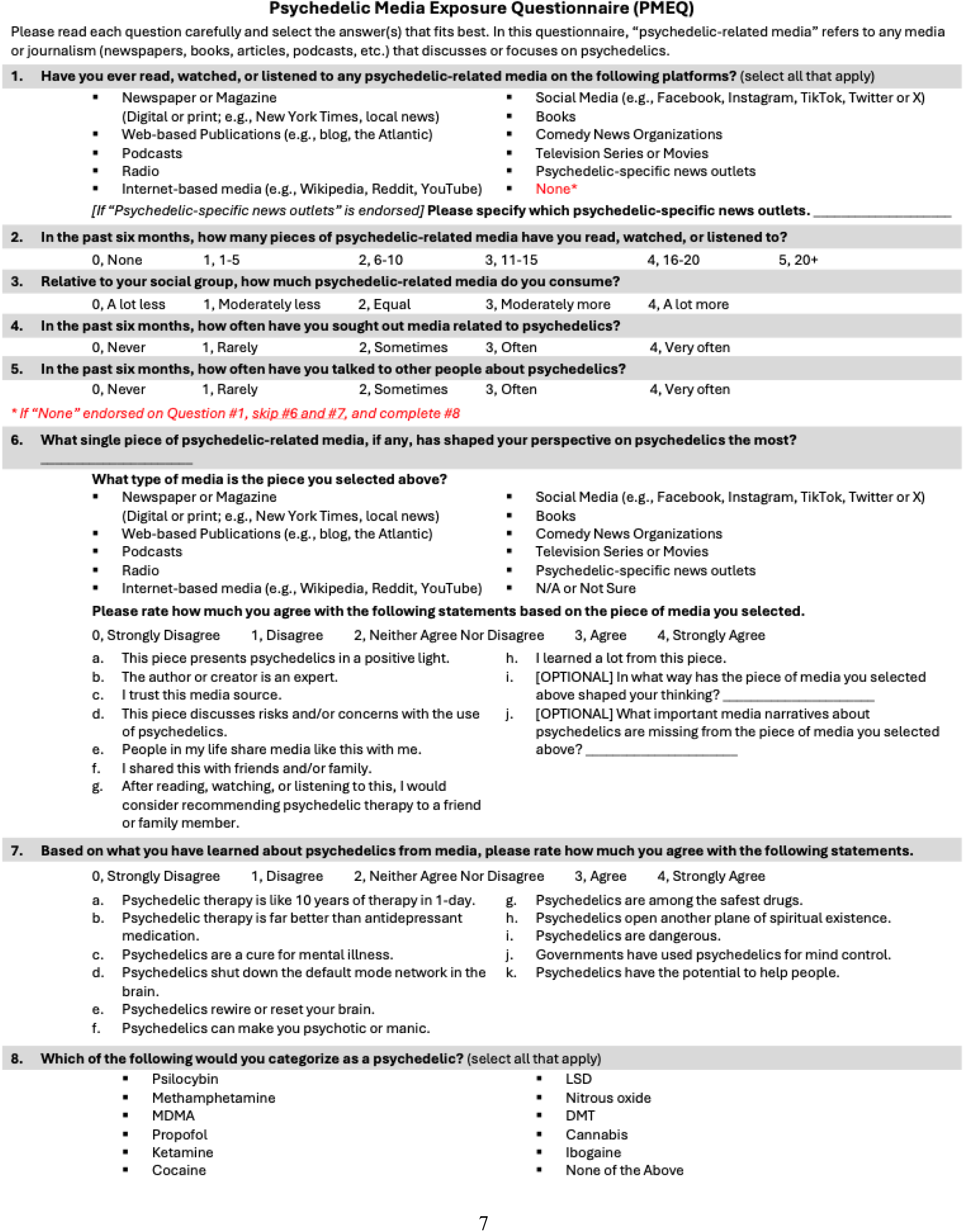
Psychedelic Media Exposure Questionnaire with scoring logic.

In the next section (Questions 7a-7k), participants are asked to rate their agreement with common phrases used in psychedelic-related media based on what they have learned about psychedelics from the media. The list of common phrases used in psychedelic-related media was pulled directly from popular media sources. These phrases are used to capture a variety of attitudes toward psychedelics in different domains: the efficacy of psychedelics for the treatment of mental illnesses, the mechanisms of change in psychedelic therapy, and the safety of psychedelics. The final question (Question 8) is included as an exploratory item to assess what participants believe to be classified as “psychedelics,” given evidence of broadly varied views about what constitutes a psychedelic drug (Žuljević et al., 2022). This item does not contribute to the score.

After item development and iteration, expert reviewers (N = 11) were recruited via the authors’ professional networks. Feedback was provided by the experts via a REDCap survey. Changes to the scale were made to improve clarity and specificity.

### Scoring

The proposed scoring structure utilizes one overall score that sums all items, reverse scoring negative attitude items, and three subscores: exposure, engagement, and attitude. The exposure subscore takes the sum of items one and two for a simple exposure score. The engagement subscore takes the sum of engagement items, defined as participants sharing, discussing, and interest in psychedelic-related media. Lastly, the attitude sub-score takes the sum of items in question seven, reverse scoring negative attitude items, for a measure of agreement with positive media “tropes” of psychedelics.

**PMEQ Score:** pmeq1 + pmeq2 + pmeq3 + pmeq4 + pmeq5 + pmeq6a + pmeq6b + pmeq6c + pmeq6d + pmeq6e + pmeq6f + pmeq6g + pmeq6h + pmeq7a + pmeq7b + pmeq7c + pmeq7d + pmeq7e + pmeq7f(*R) + pmeq7g + pmeq7h + pmeq7i(*R) + pmeq7j(*R) + pmeq7k

**Exposure Score:** pmeq1 + pmeq2

**Engagement Score:** pmeq3 + pmeq4 + pmeq5 + pmeq6e + pmeq6f + pmeq6g + pmeq6h

**Attitude Score:** pmeq7a + pmeq7b + pmeq7c + pmeq7d + pmeq7e + pmeq7f(*R) + pmeq7g + pmeq7h + pmeq7i(*R) + pmeq7j(*R) + pmeq7k

**Phase 2: Psychometric Validation**

## Methods

### Study 1: Psychometric Validation with a Stanford Medical Center Research Registry

*Recruitment*: We made use of the Stanford Pain Clinic Collaborative Health Outcomes Information Registry (CHOIR), which created and maintains a registry of Stanford Medical Center patients who had previously consented to be contacted for future research purposes (N = 6,891). The CHOIR contact registry includes patients who are recruited for multiple trials, including some who had been contacted to participate in a psilocybin (psychedelic) clinical trial for chronic low back pain (NCT05351541). The study team contacted these participants via email using an IRB-approved email template, and a portion of recipients agreed to complete the survey (N = 676). Of the participants who agreed to the consent form, a majority (N = 472) completed the survey. All 472 participants in Study 1 were analyzed for tests of reliability and validity.

*Procedures*: Participants who responded to the survey link were guided to an online information sheet about the study. All participants agreed to the terms of the study and completed the Psychedelic Media Exposure Questionnaire (PMEQ) and demographic information. See Figure 1.

*Data Analysis:* Descriptive statistics were computed for the individual items to provide an overview of the data distribution. The mean, standard deviation, skewness, and kurtosis were examined to assess the central tendency and variability of the responses. Reliability analyses were conducted to assess the internal consistency of the scale.

McDonald’s Omega (ω) and Cronbach’s alpha (*α*) coefficients were calculated to determine the extent to which items within the scale were interrelated (Hayes & Coutts, 2020). An exploratory factor analysis (EFA) was performed on the items in the first seven questions to explore the underlying structure of the scale and assess its validity. All analyses were performed using JASP version 0.18.3. To test the sample’s adequacy for EFA, we chose the Kaiser-Meyer-Olkin measure of sampling adequacy and Bartlett’s test of sphericity. Maximum likelihood factoring was chosen as the extraction method, which allows for the estimation of model fit indices and ensures consistency when moving to the confirmatory factor analysis in the second study. The number of factors to retain was decided using two methods: parallel analysis, which is robust against the overestimation of factor numbers, and visual examination of the scree plot. A Promax rotation was applied due to the anticipated correlation among factors. The factor loading cutoff was.4.

### Study 2: Psychometric Validation with Online Platforms

*Recruitment*: An IRB-approved post was repeatedly posted on online platforms, including Reddit, Facebook, and X (Twitter). A portion of users agreed to complete the survey (N = 331). Of the participants who agreed to the consent form, a portion (N = 159) completed the survey. All 159 participants in Study 2 were analyzed for tests of reliability and validity.

*Procedures*: Participants who responded to the survey link were guided to an online information sheet about the study. All participants agreed to the terms of the study and completed the Psychedelic Media Exposure Questionnaire (PMEQ) and demographic information.

*Data Analysis:* Descriptive statistics were computed for the individual items to provide an overview of the data distribution. The mean, standard deviation, skewness, and kurtosis were examined to assess the central tendency and variability of the responses. Reliability analyses were conducted to assess the internal consistency of the scale.

McDonald’s Omega (ω) and Cronbach’s alpha (*α*) coefficient were calculated to determine the extent to which items within the scale were interrelated. A confirmatory factor analysis was performed to validate the underlying structure identified through EFA in Study 1, employing Maximum Likelihood estimation because of its suitability for assessing model fit and for continuity with the EFA phase. Based on the EFA results, the hypothesized model included four latent factors with items loading on their respective factors as identified in the exploratory phase, and the factors were allowed to correlate.

## Results

### Study 1: Psychometric Validation with the Stanford University Chronic Pain Registry

#### Characteristics of Participants

Four hundred seventy-two people participated. The majority of participants were women (61.9%), over the age of 45 (81.2%), White (83.9%), had obtained or were pursuing a graduate degree (36.0%), and had a household income of over $200,000 per year (26.3%). Detailed sociodemographic characteristics are presented (**Table 1**). All data was included in the analyses for Study 1.

**Table 1.** Study 1 participant characteristics.

|  |  |
| --- | --- |
| *Age* | **N = 472** |
| 18-24 years old | 5 (1.1%) |
| 25-34 years old | 17 (3.6%) |
| 35-44 years old | 67 (14.2%) |
| 45-54 years old | 103 (21.8%) |
| 55-64 years old | 124 (26.3%) |
| 65-74 years old | 106 (22.5%) |
| 75-89 years old | 49 (10.4%) |
| 90 years or older | 1 (.2%) |
| *Gender* |  |
| Man | 170 (36.0%) |
| Woman | 292 (61.9%) |
| Non-binary | 7 (1.5%) |
| Prefer not to say | 3 (.6%) |
| Transgender | 5 (1.1%) |
| *Race* |  |
| American Indian or Alaska Native | 17 (3.6%) |
| Asian | 27 (5.7%) |
| Black or African American | 16 (3.6%) |
| Native Hawaiian or Other Pacific Islander | 2 (.4%) |
| White | 396 (83.9%) |
| Other | 44 (9.3%) |
| Hispanic or Latino | 45 (9.5%) |
| *Education Level* |  |
| Less than high school | 3 (.6%) |
| Completed high school or GED | 17 (3.6%) |
| Completed trade school | 10 (2.1%) |
| Some college | 76 (16.1%) |
| Two-year college | 33 (7%) |
| Four-year college/bachelor's degree | 121 (25.6%) |
| Some graduate school | 42 (8.9%) |
| Obtained or pursuing graduate degree | 170 (36.0%) |
| *Household Annual Income* |  |
| Less than \$10,000 | 13 (2.8%) |
| \$10,000 to \$24,999 | 40 (8.5%) |
| \$25,000 to \$49,999 | 55 (11.7%) |
| \$50,000 to \$74,999 | 53 (11.2%) |
| \$75,000 to \$99,999 | 39 (8.3%) |
| \$100,000 to \$149,999 | 93 (19.7%) |
| \$150,000 to \$199,999 | 55 (11.7%) |
| \$200,000 and above | 124 (26.3%) |

#### PMEQ Descriptive Statistics

The mean total score on the PMEQ for Study 1 was M=42.88 (SD=22.58), with a range of 0-85. Mean scores on subscales were as follows: *Engagement* M = 14.05 (SD=5.14, Range: 0-28), *Exposure* M = 4.88 (SD=3.08, Range: 0-13), *Attitude* M=23.66 (SD=5.98, Range 3-42). All items in the PMEQ were utilized to their full range. Descriptive statistics for each individual item are presented in **Table 2**.

**Table 2.** Study 1 Descriptive Statistics. * Indicates items are reverse scored.

| Item | Range | Mean | SD | Skewness | Kurtosis |
| --- | --- | --- | --- | --- | --- |
| Item 1 | 0-9 | 2.41 | 2.121 | 0.895 | 0.216 |
| Item 2 | 0-5 | 2.47 | 1.927 | 0.352 | -1.58 |
| Item 3 | 0-4 | 1.84 | 1.307 | -0.156 | -1.145 |
| Item 4 | 0-4 | 0.97 | 1.035 | 0.817 | -0.012 |
| Item 5 | 0-4 | 1.33 | 1.052 | 0.48 | -0.267 |
| Item 6a | 0-4 | 3.01 | 0.824 | -0.586 | 0.467 |
| Item 6b | 0-4 | 2.58 | 0.933 | -0.05 | -0.437 |
| Item 6c | 0-4 | 2.97 | 0.844 | -0.534 | 0.116 |
| Item 6d* | 0-4 | 1.22 | 0.901 | 0.818 | 0.668 |
| Item 6e | 0-4 | 1.89 | 1.242 | -0.106 | -1.135 |
| Item 6f | 0-4 | 1.96 | 1.287 | -0.177 | -1.204 |
| Item 6g | 0-4 | 2.5 | 1.137 | -0.542 | -0.343 |
| Item 6h | 0-4 | 2.83 | 0.919 | -0.685 | 0.312 |
| Item 7a | 0-4 | 1.88 | 1.026 | -0.056 | -0.154 |
| Item 7b | 0-4 | 2.25 | 0.999 | -0.12 | 0.092 |
| Item 7c | 0-4 | 1.47 | 1.018 | 0.16 | -0.423 |
| Item 7d | 0-4 | 2.09 | 0.757 | -0.185 | 1.665 |
| Item 7e | 0-4 | 2.34 | 0.868 | -0.338 | 0.247 |
| Item 7f* | 0-4 | 2.00 | 1.015 | 0.237 | -0.404 |
| Item 7g | 0-4 | 1.74 | 0.913 | -0.087 | 0.106 |
| Item 7h | 0-4 | 2.41 | 1.028 | -0.598 | 0.158 |
| Item 7i* | 0-4 | 2.14 | 0.93 | -0.024 | 0.099 |
| Item 7j* | 0-4 | 1.98 | 1.142 | 0.313 | -0.543 |
| Item 7k | 0-4 | 3.35 | .712 | -1.172 | 2.286 |

#### Additional Analyses

Due to the older sample population compared to results in large randomized-controlled trials (Goodwin et al., 2022), we assessed bias related to age. Bivariate correlation was conducted to examine the relationship between PMEQ score and age. Non-parametric correlation was conducted due to the categorical measurement of age. Age and PMEQ score were significantly negatively correlated (ρ=-.193, p<.001).

#### Reliability

McDonald’s Omega and Cronbach’s alpha analyses indicate good internal consistency (ω =.83 and *α* =.84).

#### Exploratory Factor Analysis

The Kaiser-Meyer-Olkin (KMO) measure of sampling adequacy was 0.87, and Bartlett’s test of sphericity was significant (χ²(276) = 3861.85, *p* <.001), indicating the data were suitable for factor analysis.

Parallel analysis of the items from the first seven questions yielded a four-factor solution (*λ_1_=6.54, λ_2_=2.17, λ_3_=1.84, λ_4_=1.51)*, which together explained 41% of the variance in responses in line with standard sociometric tools (Wood et al., 2015). Retention of four factors was supported by a visual examination of the scree plot, where an elbow was noted after the fourth factor.

The factor loadings after Promax rotation revealed that one item had loadings >0.4 on more than one factor. This item, “Psychedelics are among the safest drugs” (7g), was designed to reflect media-informed attitudes regarding the safety of psychedelics. In this analysis, 7g loads on both positive and negative media-informed attitude factors, which was theorized. Despite the cross-loading of the item, the model fit indices were in the acceptable range (e.g., CFI=.91, root mean square of residuals or RMSEA=.06), and the internal consistency of the individual factors was acceptable (Factor 1=.82, Factor 2=.76, Factor 3=.73, Factor 4=.66).

Factor 1 was primarily associated with a positive media-informed attitude toward psychedelics, Factor 2 with subjective assessment of an emblematic, attitude-informing piece, Factor 3 with engagement with psychedelics-related media sources, and Factor 4 with a negative media-informed attitude toward psychedelics reflecting risks of the compounds. The factors were moderately correlated, as expected. Of note, items with low communalities (below 0.5) were reviewed for potential revision or removal but were ultimately retained, as they provided unique and valuable insights.

The results of the exploratory factor analysis supported a four-factor solution for this newly designed scale for psychedelic-related media exposure and media-informed attitudes.

#### Exploratory Analysis

Participants reported what they believed to be classified as a “psychedelic” in the final question of the measure. Results (**Table 3**) indicate that the majority of participants identified psilocybin (77.5%) and LSD (85.8%) as psychedelics, followed by MDMA (43.4%) and ketamine (41.7%).

**Table 3.** Study 1 responses to “Which of the following would you categorize as a psychedelic?”

| **Drug** | **N (%)** |
| --- | --- |
| Psilocybin | 366 (77.5%) |
| Methamphetamine | 38 (8.1%) |
| MDMA | 205 (43.4%) |
| Propofol | 26 (5.5%) |
| Ketamine | 197 (41.7%) |
| Cocaine | 29 (6.1%) |
| LSD | 405 (85.8%) |
| Nitrous Oxide | 38 (8.1%) |
| DMT | 128 (27.1%) |
| Cannabis | 104 (22%) |
| Ibogaine | 69 (14.6%) |

After completion of Study 1, minor revisions to the measure were made to improve clarity and factor cohesion based on participant feedback. Further, the demographic characteristics of Study 1 were predominantly highly educated, wealthy, white women, so Study 2 aimed to target a different demographic sample of online platform users.

### Study 2: Psychometric Validation with Online Platforms

#### Characteristics of Participants

One hundred fifty-nine people participated. The majority of participants were men (54%), 25-35 years old (40.9%), White (83%), had a bachelor’s degree (34.6%), and had a household income of $25,000 to $49,999 per year (19.5%). Detailed sociodemographic characteristics are presented (**Table 4**). All data was included in the analyses for Study 2.

**Table 4.** Study 2 Participant characteristics.

|  |  |
| --- | --- |
| *Age* | N = 159 |
| 18-24 years old | 50 (31.4%) |
| 25-34 years old | 65 (40.9%) |
| 35-44 years old | 28 (17.6%) |
| 45-54 years old | 11 (6.9%) |
| 55-64 years old | 3 (1.9%) |
| 65-74 years old | 2 (1.3%) |
| *Gender* |  |
| Man | 86 (54%) |
| Woman | 63 (39.6%) |
| Non-binary | 6 (3.8%) |
| Prefer not to say | 4 (2.5%) |
| Transgender | 13 (8.2%) |
| *Race* |  |
| American Indian or Alaska Native | 2 (1.3%) |
| Asian | 19 (11.9%) |
| Black or African American | 4 (2.5%) |
| Native Hawaiian or Other Pacific Islander | 3 (1.9%) |
| White | 132 (83%) |
| Other | 14 (8.8%) |
| Hispanic or Latino | 10 (6.3%) |
| *Education Level* |  |
| Less than high school | 2 (1.3%) |
| Completed high school or GED | 6 (3.8%) |
| Completed trade school | 1 (.6%) |
| Some college | 25 (15.7%) |
| Two-year college | 8 (5%) |
| Four-year college/bachelor's degree | 55 (34.6%) |
| Some graduate school | 10 (6.3%) |
| Obtained or pursuing graduate degree | 52 (32.7%) |
| *Household Annual Income* |  |
| Less than \$10,000 | 12 (7.5%) |
| \$10,000 to \$24,999 | 17 (10.7%) |
| \$25,000 to \$49,999 | 31 (19.5%) |
| \$50,000 to \$74,999 | 23 (14.5%) |
| \$75,000 to \$99,999 | 15 (9.4%) |
| \$100,000 to \$149,999 | 28 (17.6%) |
| \$150,000 to \$199,999 | 9 (5.7%) |
| \$200,000 and above | 24 (15.1%) |

#### PMEQ Descriptive Statistics

The mean total score on the PMEQ for Study 2 was M=47.83 (SD=22.31), with a range of 0-89. Mean scores on subscales were as follows: *Engagement* M = 15.42 (SD=5.72, Range: 1-28), *Exposure* M = 6.34 (SD=4.38, Range: 0-15), *Attitude* M=27.06 (SD=6.49, Range 8-43). All items in the PMEQ were utilized to their full range, excluding the last item, “psychedelics have the potential to help people.” Descriptive statistics for each individual item are presented in **Table 5**.

**Table 5.**
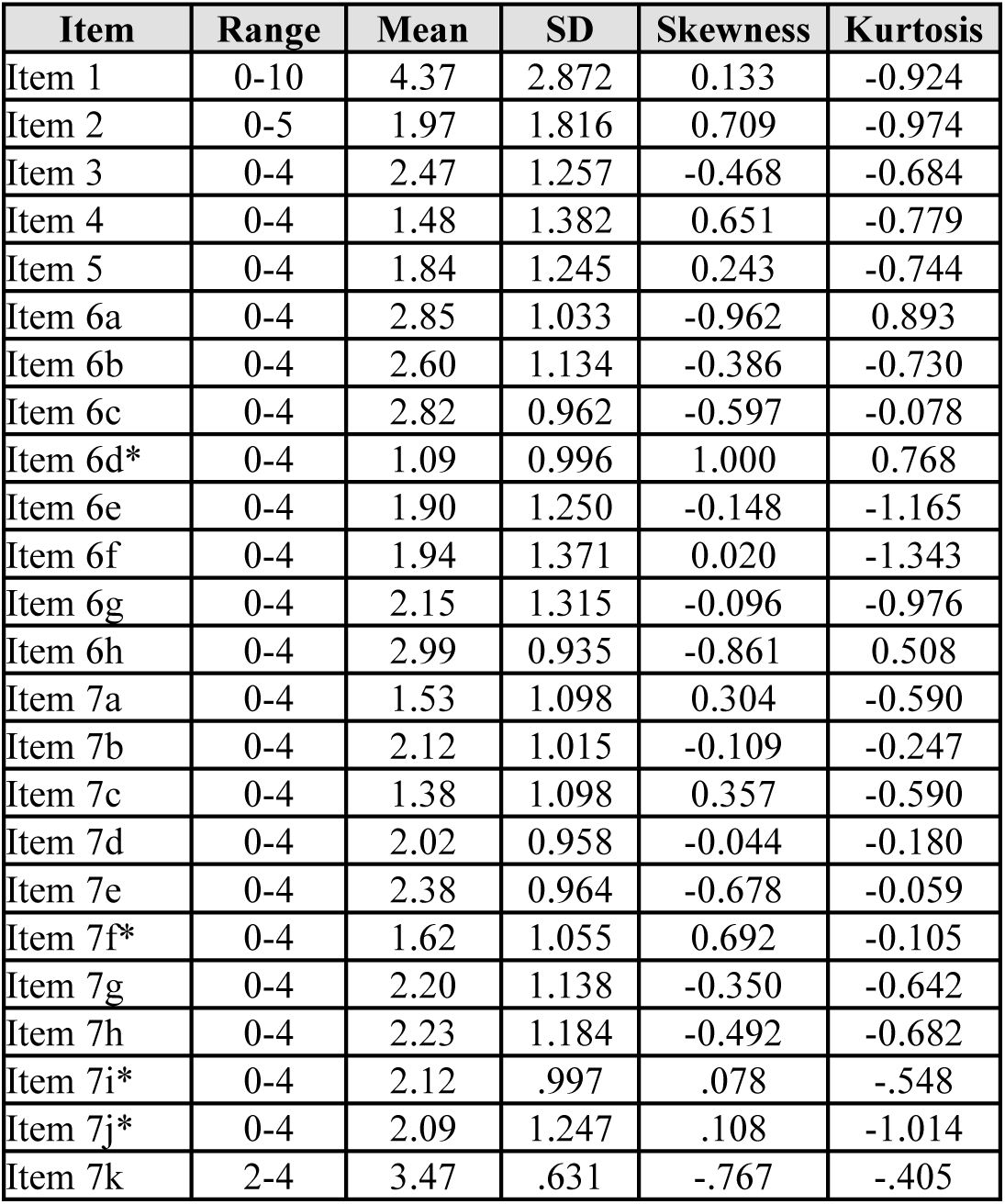
Study 2 Descriptive Statistics. * Indicates items are reverse scored.

| Item | Range | Mean | SD | Skewness | Kurtosis |
| --- | --- | --- | --- | --- | --- |
| Item 1 | 0-10 | 4.37 | 2.872 | 0.133 | -0.924 |
| Item 2 | 0-5 | 1.97 | 1.816 | 0.709 | -0.974 |
| Item 3 | 0-4 | 2.47 | 1.257 | -0.468 | -0.684 |
| Item 4 | 0-4 | 1.48 | 1.382 | 0.651 | -0.779 |
| Item 5 | 0-4 | 1.84 | 1.245 | 0.243 | -0.744 |
| Item 6a | 0-4 | 2.85 | 1.033 | -0.962 | 0.893 |
| Item 6b | 0-4 | 2.60 | 1.134 | -0.386 | -0.730 |
| Item 6c | 0-4 | 2.82 | 0.962 | -0.597 | -0.078 |
| Item 6d* | 0-4 | 1.09 | 0.996 | 1.000 | 0.768 |
| Item 6e | 0-4 | 1.90 | 1.250 | -0.148 | -1.165 |
| Item 6f | 0-4 | 1.94 | 1.371 | 0.020 | -1.343 |
| Item 6g | 0-4 | 2.15 | 1.315 | -0.096 | -0.976 |
| Item 6h | 0-4 | 2.99 | 0.935 | -0.861 | 0.508 |
| Item 7a | 0-4 | 1.53 | 1.098 | 0.304 | -0.590 |
| Item 7b | 0-4 | 2.12 | 1.015 | -0.109 | -0.247 |
| Item 7c | 0-4 | 1.38 | 1.098 | 0.357 | -0.590 |
| Item 7d | 0-4 | 2.02 | 0.958 | -0.044 | -0.180 |
| Item 7e | 0-4 | 2.38 | 0.964 | -0.678 | -0.059 |
| Item 7f* | 0-4 | 1.62 | 1.055 | 0.692 | -0.105 |
| Item 7g | 0-4 | 2.20 | 1.138 | -0.350 | -0.642 |
| Item 7h | 0-4 | 2.23 | 1.184 | -0.492 | -0.682 |
| Item 7i* | 0-4 | 2.12 | .997 | .078 | -.548 |
| Item 7j* | 0-4 | 2.09 | 1.247 | .108 | -1.014 |
| Item 7k | 2-4 | 3.47 | .631 | -.767 | -.405 |

#### Additional Analyses

In the second, younger sample, we again sought to identify a relationship between PMEQ score and age. Bivariate correlation was conducted to examine the relationship between PMEQ score and age. Non-parametric correlation was conducted due to the categorical measurement of age. Age and PMEQ score were significantly positively correlated (ρ=.250, p=.001).

#### Reliability

McDonald’s Omega and Cronbach’s alpha analyses indicate good internal consistency (*ω* =.86 and *α* =.85).

#### Confirmatory Factor Analysis

Fit indices indicated an acceptable model fit: comparative fit index (CFI) =.94, Tucker-Lewis index =.93, root mean square error of approximation (RMSEA) =.05, and standardized root mean square residual (SRMR) =.07. Factors analysis results are visualized in **Figure 2**. The standardized factor loadings ranged from.33 to 2.20, indicating that items had substantial loadings on their respective factors, and R-squared values ranged from.18 to 1.00, demonstrating that the factors explained a substantial proportion of variance in most of the observed variables. The correlations between the factors ranged from.30 to.58, indicating moderate relationships among the factors. All the residual variances were significant (p <.001), with several indicators (e.g., item 1, item 7h) notably high, indicating that there is unexplained variance in these indicators.

**Figure 2.**
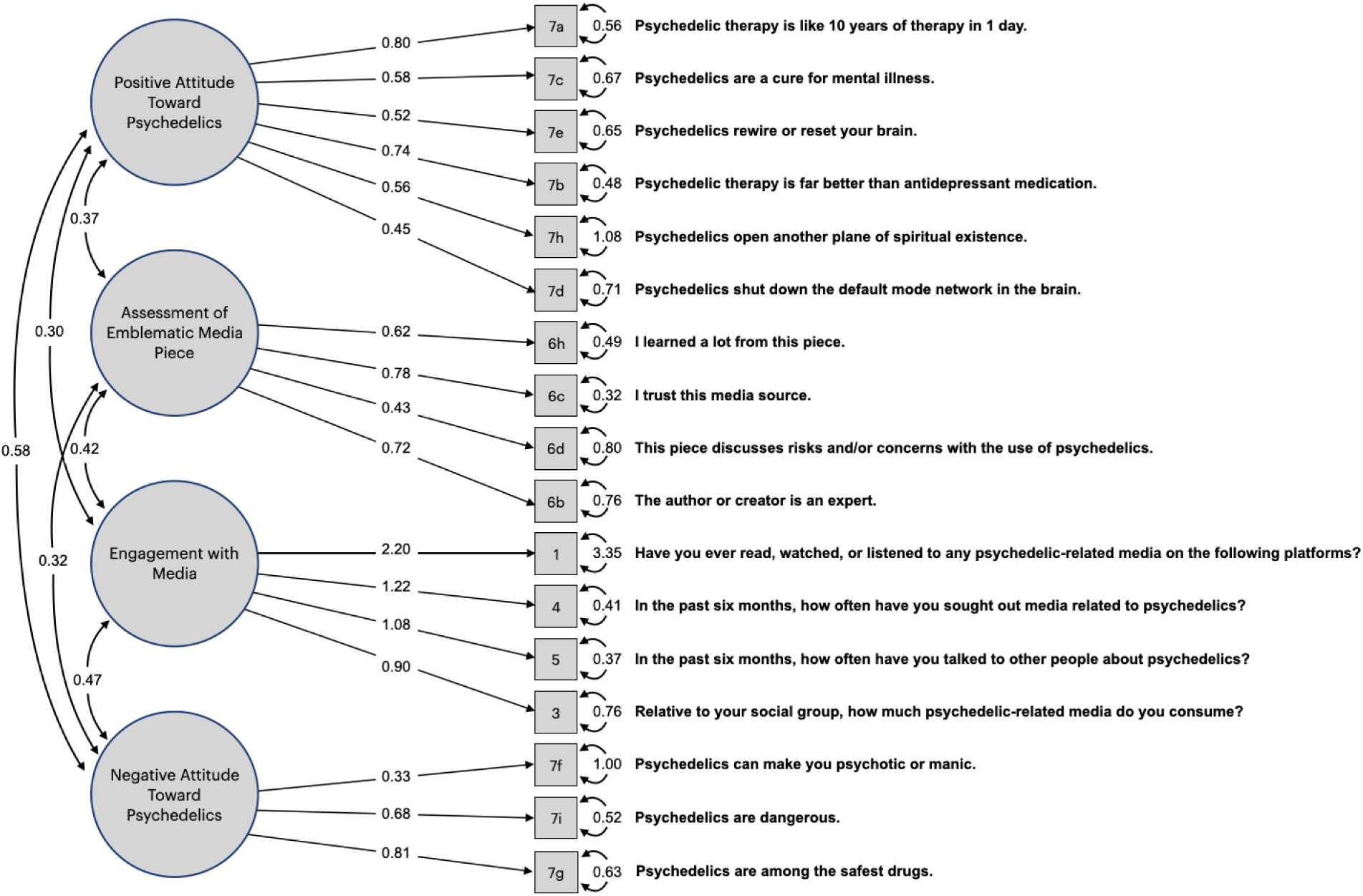
Confirmatory factor analysis results. Factors (circled) connect to each other by curved arrows and associated correlation between the factors; and connect to individual questionnaire items by straight arrows and the associated factor loading. Numbers immediately right of boxed item numbers indicate residual variance.

#### Exploratory Analysis

Participants reported what they believed to be classified as a “psychedelic” in the final question of the measure. Results (**Table 6**) indicate that the majority of participants identified psilocybin (85.5%), MDMA (57.2%), LSD (93.7%), and DMT (72.3%) as psychedelics, followed by ketamine (47.8%) and Ibogaine (29.6%).

**Table 6.** Study 2 responses to “Which of the following would you categorize as a psychedelic?”

| **Drug** | **N (%)** |
| --- | --- |
| Psilocybin | 136 (85.5%) |
| Methamphetamine | 14 (8.8%) |
| MDMA | 91 (57.2%) |
| Propofol | 8 (5%) |
| Ketamine | 76 (47.8%) |
| Cocaine | 9 (5.7%) |
| LSD | 149 (93.7%) |
| Nitrous Oxide | 15 (9.4%) |
| DMT | 115 (72.3%) |
| Cannabis | 40 (25.2%) |
| Ibogaine | 47 (29.6%) |
| None of the Above | 1 (.6%) |

Results from the confirmatory factor analysis generally support the four-factor model identified in the exploratory factor analysis in Study 1, although several indicators exhibited high residual variances, particularly the items related to total exposure to psychedelic-related media (item 1) and the belief that psychedelics open another plane of spiritual existence (item 7h), suggesting substantial unexplained variance. Item 1 is an assessment of the respondent’s subjective recall of exposure to psychedelic-related media over the past 6 months, which was hypothesized to potentially, but not completely, influence attitudes related to psychedelics. Retaining these items reflects the intended purpose of the scale, the goal of which is to capture exposure to common media tropes at a critical juncture of increased clinical research and media attention.

## Discussion

The present study describes the development and initial psychometric properties of a new scale, the PMEQ, which measures exposure to psychedelic-related media and attitudes based on exposure. The results of our analyses support the validation of the PMEQ as an instrument to measure the construct of psychedelic-related media exposure, which demonstrated good internal consistency, with four underlying factors (positive attitude toward psychedelics, assessment of emblematic media piece, engagement with media, and negative attitude toward psychedelics). The PMEQ was validated in a three-step process: 1) item generation and consultation, 2) administration of the measure to patients in a clinical research registry, and 3) administration of the measure to a sample of participants recruited on social media platforms.

The factor structure of the PMEQ indicated that a positive attitude is inherent in psychedelic-related media exposure. This finding aligns with the anecdotal sentiment that psychedelic-related media is largely positive (Aday et al., 2022; Meling et al., 2024; Yaden et al., 2022). However, in August of 2024, after the period of data collection for this study, the FDA rejected Lykos Therapeutic’s application for MDMA for the treatment of PTSD (Mahase, 2024). Media and public sentiment toward psychedelics may be in the midst of a shift, in ways that echo the shift in the period of the 1960s (Abraham et al., 1996; Noorani, 2020) and may come to be recursively reflected in subsequent participant and patient accounts.

Our exploratory analyses of the relationship between age and PMEQ score sampled across two populations (older age in study one and younger age in study two) supports a peak in media exposure in the 30-40 year old range, with declining media exposure on both ends of the age spectrum.

Several items emerged as particularly distinct and did not align well with the four-factor structures identified in the initial study and explored in the subsequent study. Notably, these items pertained to the perceived dangers of psychedelics, their association with spirituality, and their potential to induce manic or psychotic states. These items were intentionally retained in the scale and reviewed with the panel of experts to capture current perceptions of psychedelics. Historically, such tropes were prevalent, particularly during the first few decades of research in psychedelics. Although these perceptions remain to some extent, as is revealed in media representations, there has been a noticeable shift toward the medicalization of psychedelics. In both the research registry sample and online sample, participants endorsed intermediate scores within the full scale range and at the subscore level, suggesting an appropriate sampling of the population variance. LSD, psilocybin, DMT, and MDMA were the most widely recognized psychedelics.

### Strengths & Limitations

One strength of our study is that the CHOIR registry captured a sample of patients whose condition, chronic pain, is being investigated for treatment with psilocybin. This registry has been previously used for email recruitment for a trial of psilocybin therapy (NCT05351541), which is precisely the population of interest for assessing potential psychedelic-related media exposure and expectancy bias. However, we note that the majority of clinical trials of psilocybin and MDMA are intended for patients with treatment-resistant depression and posttraumatic stress disorder, respectively, and may not have the same media exposure or engagement as patients with chronic low back pain.

The primary limitation of our study was the risk of selection bias, as is common with survey studies. We attempted to sample a diverse sample utilizing the CHOIR registry, however we obtained a largely white, wealthy, female participant pool. We emphasized that no prior experience or knowledge of psychedelics was necessary to participate, but there likely remained a participation bias for those who were interested in the subject. The second study utilized social media platforms (primarily Reddit and X/Twitter) to recruit a younger male population. Our sample nonetheless remained predominantly white. Therefore, the PMEQ may be limited in its applicability in racially diverse samples. Additionally, we cannot claim that our initial item pool for the PMEQ was fully comprehensive, as there may be other unknown aspects of media exposure that were not captured through our instrument.

### Future research

We recommend the use of the PMEQ in clinical research studies of psychedelics to account for exposure to positive media reports on psychedelics that may be contributing to expectations for treatment. Expectations are complex constructs to be measured. While existing measures, such as the Stanford Expectations of Treatment Scale (SETS; Younger et al., 2012), may be useful in this endeavor, there is likely a need for a psychedelic-specific expectation scale (Gukasyan & Nayak, 2022). The PMEQ partially fulfills this need and highlights the multifaceted nature of measuring expectations. The psychological support model of many psychedelic clinical trials could incorporate the PMEQ as the basis for a study therapist to discuss expectations and media exposure that trial participants may have coming into the study. Provider expectations of treatment success may be similarly influenced by media expectations and could be measured with the PMEQ to assess a “socially transmitted placebo effect” (Chen et al., 2019). We anticipate that future iterations of this scale may update the referenced media-informed beliefs and explore the relationship between clinical research populations and the more diverse general population.

This study demonstrated the reliability and validity of the PMEQ in assessing exposure to media related to psychedelics, which is increasingly important as there is more media attention and accelerating clinical research using psychedelics. Further, given masking challenges and potential expectancy effects, we encourage the replication and exploration of our new instrument in different settings and populations to best account for these factors.

## Data Availability

The data that support the findings of this study are available from the co-corresponding authors (AE and BDH) upon reasonable request.

## Author Contributions

- **Conceptualization and Study Design**: AE, CK, SL and BDH conceived the study, formulated the research hypotheses, and designed the overall study framework including obtaining regulatory approval.
- **Scale Development and Validation:** AE, CK, TN, SL and BDH developed the new scale—including item generation, pilot testing, and establishing preliminary psychometric properties—and planned the validation procedures.
- **Data Collection**: AE and CK managed participant recruitment and oversaw the collection of clinical trial data.
- **Data Analysis and Interpretation:** AE and CK performed the statistical analyses, interpreted the findings, and ensured that the data supported the scale’s validity.
- **Manuscript Drafting:** AE, CK and BDH prepared the initial draft of the manuscript.
- **Manuscript Review and Editing:** AE, CK, TN, SL and BDH critically revised the manuscript for content and clarity.
- **Final Approval:** All authors have read and approved the final version of the manuscript.

## References

Abraham, H. D., Abridge, A. M., & Gogig, P. (1996). The Psychopharmacology of Hallucinogens. Neuropsychopharmacology, 14, 285–298.

Aday, J. S., Heifets, B. D., Pratscher, S. D., Bradley, E., Rosen, R., & Woolley, J. D. (2022). Great Expectations: Recommendations for improving the methodological rigor of psychedelic clinical trials. Psychopharmacology, 239(6), 1989–2010. 10.1007/s00213-022-06123-7

Carpenter, D. (2020). 5-MeO-DMT: The 20-Minute Psychoactive Toad Experience That’s Transforming Lives. https://www.forbes.com/sites/davidcarpenter/2020/02/02/5-meo-dmt-the-20-minute-psychoactive-toad-experience-thats-transforming-lives/

Chen, P.-H. A., Cheong, J. H., Jolly, E., Elhence, H., Wager, T. D., & Chang, L. J. (2019). Socially transmitted placebo effects. Nature Human Behaviour, 3(12), 1295–1305. 10.1038/s41562-019-0749-5

Goodwin, G. M., Aaronson, S. T., Alvarez, O., Arden, P. C., Baker, A., Bennett, J. C., Bird, C., Blom, R. E., Brennan, C., Brusch, D., Burke, L., Campbell-Coker, K., Carhart-Harris, R., Cattell, J., Daniel, A., DeBattista, C., Dunlop, B. W., Eisen, K., Feifel, D.,… Malievskaia, E. (2022). Single-Dose Psilocybin for a Treatment-Resistant Episode of Major Depression. New England Journal of Medicine, 387(18), 1637– 1648. 10.1056/NEJMoa2206443

Gukasyan, N., & Nayak, S. M. (2022). Psychedelics, placebo effects, and set and setting: Insights from common factors theory of psychotherapy. Transcultural Psychiatry, 59(5), 652–664. 10.1177/1363461520983684

Hall, W. D., & Humphreys, K. (2022). Is good science leading the way in the therapeutic use of psychedelic drugs? Psychological Medicine, 52(14), 2849–2851. 10.1017/S0033291722003191

Hayes, A. F., & Coutts, J. J. (2020). Use Omega Rather than Cronbach’s Alpha for Estimating Reliability. But…. Communication Methods and Measures, 14(1), 1–24. 10.1080/19312458.2020.1718629

Kamenova, K., & Caulfield, T. (2015). Stem cell hype: Media portrayal of therapy translation. Science Translational Medicine, 7(278). 10.1126/scitranslmed.3010496

Kashdan, T. B., Disabato, D. J., Goodman, F. R., Doorley, J. D., & McKnight, P. E. (2020). Understanding psychological flexibility: A multimethod exploration of pursuing valued goals despite the presence of distress. Psychological Assessment, 32(9), 829–850. 10.1037/pas0000834

Love, S. (2023). Scientists Gave People Psychedelics—And Then Erased Their Memory | WIRED. https://www.wired.com/story/psychedelics-study-design-research-rcts/

Luoma, J. B., Chwyl, C., Bathje, G. J., Davis, A. K., & Lancelotta, R. (2020). A Meta-Analysis of Placebo-Controlled Trials of Psychedelic-Assisted Therapy. Journal of Psychoactive Drugs, 52(4), 289–299. 10.1080/02791072.2020.1769878

Mahase, E. (2024). MDMA assisted therapy: Three papers are retracted as FDA rejects PTSD application. *BMJ*, q1798. 10.1136/bmj.q1798

Mameli, F., Zirone, E., Girlando, R., Scagliotti, E., Rigamonti, G., Aiello, E. N., Poletti, B., Ferrucci, R., Ticozzi, N., Silani, V., Locatelli, M., Barbieri, S., & Ruggiero, F. (2023). Role of expectations in clinical outcomes after deep brain stimulation in patients with Parkinson’s disease: A systematic review. Journal of Neurology, 270(11), 5274–5287. 10.1007/s00415-023-11898-6

Meling, D., Ehrenkranz, R., Nayak, S. M., Aicher, H. D., Funk, X., Van Elk, M., Graziosi, M., Bauer, P. R., Scheidegger, M., & Yaden, D. B. (2024). Mind the Psychedelic Hype: Characterizing the Risks and Benefits of Psychedelics for Depression. Psychoactives, 3(2), 215–234. 10.3390/psychoactives3020014

Mitchell, J. M., & Anderson, B. T. (2024). Psychedelic therapies reconsidered: Compounds, clinical indications, and cautious optimism. Neuropsychopharmacology, 49(1), 96–103. 10.1038/s41386-023-01656-7

Noorani, T. (2020, July 21). The Pollan Effect: Psychedelic Research between World and Word. Society for Cultural Anthropology. https://culanth.org/fieldsights/the-pollan-effect-psychedelic-research-between-world-and-word

Noorani, T., Bedi, G., & Muthukumaraswamy, S. (2023). Dark loops: Contagion effects, consistency and chemosocial matrices in psychedelic-assisted therapy trials. Psychological Medicine, 53(13), 5892–5901. 10.1017/S0033291723001289

Pollan, M. (2018). How to change your mind: What the new science of psychedelics teaches us about consciousness, dying, addiction, depression, and transcendence. Penguin Press.

Reiff, C. M., Richman, E. E., Nemeroff, C. B., Carpenter, L. L., Widge, A. S., Rodriguez, C. I., Kalin, N. H., McDonald, W. M., & the Work Group on Biomarkers and Novel Treatments, a Division of the American Psychiatric Association Council of Research. (2021). Psychedelics and Psychedelic-Assisted Psychotherapy. FOCUS, 19(1), 95–115. 10.1176/appi.focus.19104

Robledo, I., & Jankovic, J. (2017). Media hype: Patient and scientific perspectives on misleading medical news. Movement Disorders, 32(9), 1319–1323. 10.1002/mds.26993

Schatzberg, A. F. (2020). Some Comments on Psychedelic Research. American Journal of Psychiatry, 177(5), 368–369. 10.1176/appi.ajp.2020.20030272

Szigeti, B., & Heifets, B. D. (2024). Expectancy Effects in Psychedelic Trials. Biological Psychiatry. Cognitive Neuroscience and Neuroimaging, 9(5), 512–521. 10.1016/j.bpsc.2024.02.004

Szigeti, B., Weiss, B., Rosas, F. E., Erritzoe, D., Nutt, D., & Carhart-Harris, R. (2024). Assessing expectancy and suggestibility in a trial of escitalopram v. Psilocybin for depression. Psychological Medicine, 1–8. 10.1017/S0033291723003653

van Elk, M., & Fried, E. I. (2023). History repeating: Guidelines to address common problems in psychedelic science. Therapeutic Advances in Psychopharmacology, 13, 20451253231198466. 10.1177/20451253231198466

Walentynowicz, M., Schneider, S., & Stone, A. A. (2018). The effects of time frames on self-report. PLOS ONE, 13(8), e0201655. 10.1371/journal.pone.0201655

Wood, N. D., Akloubou Gnonhosou, D. C., & Bowling, J. (2015). Combining Parallel and Exploratory Factor Analysis in Identifying Relationship Scales in Secondary Data. Marriage & Family Review, 51(5), 385–395. 10.1080/01494929.2015.1059785

Yaden, D. B., Potash, J. B., & Griffiths, R. R. (2022). Preparing for the Bursting of the Psychedelic Hype Bubble. JAMA Psychiatry, 79(10), 943. 10.1001/jamapsychiatry.2022.2546

Younger, J., Gandhi, V., Hubbard, E., & Mackey, S. (2012). Development of the Stanford Expectations of Treatment Scale (SETS): A tool for measuring patient outcome expectancy in clinical trials. Clinical Trials, 9(6), 767–776. 10.1177/1740774512465064

Žuljević, M. F., Buljan, I., Leskur, M., Kaliterna, M., Hren, D., & Duplančić, D. (2022). Validation of a new instrument for assessing attitudes on psychedelics in the general population. Scientific Reports, 12(1), 18225. 10.1038/s41598-022-23056-5

